# Implementation of point-of-care screening for *Chlamydia trachomatis, Neisseria gonorrhoeae*, and *Trichomonas vaginalis* among pregnant women in South Africa: a mixed-methods process evaluation of the Philani Ndiphile trial

**DOI:** 10.64898/2026.04.08.26350414

**Authors:** Natalie G. Shaetonhodi, Lindsey De Vos, Chibuzor Babalola, Alex de Voux, Dvora Joseph Davey, Mandisa Mdingi, Remco P.H. Peters, Jeffrey D. Klausner, Andrew Medina-Marino

## Abstract

**Background:** Curable sexually transmitted infections (STIs), including *Chlamydia trachomatis, Neisseria gonorrhoeae*, and *Trichomonas vaginalis*, remain highly prevalent among pregnant women in South Africa. Despite poor diagnostic performance in pregnancy, syndromic management remains standard care. Point-of-care (POC) screening enables aetiological diagnosis and same-visit treatment but is not yet included in national guidelines. We conducted a mixed-methods process evaluation to examine determinants of antenatal POC STI screening implementation in public facilities.

**Methods:** This evaluation was embedded within the three-arm Philani Ndiphile randomized trial (March 2021-February 2025) across four public clinics in the Eastern Cape. Screening used a near-POC, electricity-dependent nucleic acid amplification test with a 90-minute turnaround time. Reach, Adoption, Implementation, and Maintenance were assessed using the RE-AIM framework. Quantitative indicators included uptake of screening, treatment, and follow-up attendance. Qualitative data included in-depth interviews with 20 pregnant women and five focus group discussions with 21 research staff and government healthcare workers. The Consolidated Framework for Implementation Research guided qualitative analysis. Findings were integrated using narrative weaving.

**Results:** Screening uptake was high (99.0%), with treatment coverage of 95.2% at baseline and 93.5% at repeat screening. Same-day treatment was lower (50.7% and 69.8%) and varied substantially by facility, reflecting operational constraints including turnaround time, patient volume, infrastructure, and electricity. Attendance was higher when screening was integrated into routine ANC. Women valued screening for infant health, while providers recognised advantages over syndromic management but highlighted workforce, resource, and maintenance constraints. Socioeconomic factors, including transport costs, hunger, and work commitments, influenced retention and waiting.

**Conclusions:** Antenatal POC STI screening was acceptable and achieved high treatment coverage in a research setting. However, same-day treatment was constrained by operational requirements of the testing platform. Scale-up will require workflow integration, strengthened health system capacity, and faster diagnostics suited to routine antenatal care.

**Key Messages:** *What is already known on this topic:* Syndromic management remains standard antenatal care in many low-resource settings despite failing to capture up to 89% of infections that remain asymptomatic. Point-of-care aetiological screening has demonstrated feasibility, acceptability, and potential clinical benefit in research settings, yet has not been widely adopted into national policy. Limited evidence exists on the health system requirements and contextual determinants influencing scale-up within routine public facilities.

*What this study adds:* This mixed-methods process evaluation demonstrates high uptake and treatment coverage of antenatal POC STI screening in a trial setting, while identifying facility-level, structural, and socioeconomic factors shaping same-day treatment and retention. We show that implementation success varies substantially across clinics and depends on assay characteristics, workflow integration, human resources, infrastructure reliability, and follow-up capacity.

*How this study might affect research, practice or policy:* These findings provide implementation-relevant evidence to inform national policy deliberations on integrating POC STI screening into antenatal care. Sustainable scale-up will require context-adapted delivery models, strengthened workforce and supply systems, faster diagnostics, and alignment with existing ANC workflows to ensure equitable and durable impact.

## Background

Curable sexually transmitted infections (STIs), including *Chlamydia (C.) trachomatis, Neisseria (N.) gonorrhoeae, and Trichomonas (T.) vaginalis*, remain highly prevalent among pregnant women in sub-Saharan Africa(1) and are associated with adverse maternal and neonatal outcomes(2–4), including increased risk of HIV acquisition and vertical transmission.(5,6) South African-based studies estimate 50–80% of curable STIs among pregnant women are asymptomatic while 40–60% presenting with STI-related symptoms test negative.(7,8) Despite these limitations, syndromic management remains standard-of-care in antenatal care (ANC) in many low-resource settings, including South Africa(9), where it is associated with untreated asymptomatic infections and unnecessary antibiotic use.(10–12)

Point-of-care (POC) screening enables aetiological diagnosis and same-visit treatment for curable STIs, offering an alternative to syndromic management and potentially reducing adverse birth outcomes,(3,13) and vertical HIV transmission.(14) High feasibility and acceptability of antenatal POC STI screening for *C. trachomatis, N. gonorrhoeae*, and *T. vaginalis* have been demonstrated in South Africa,(15) and although decentralised diagnostic platforms for STIs are included in the national strategic plan (2023–2028),(16) they have not yet been incorporated into national testing guidelines or clinical algorithms. Across Africa, a persistent gap between evidence and policy reflects competing priorities and limited understanding of system requirements for scale-up in resource-constrained settings.(17–19) Characterising implementation processes is therefore critical for informing policy and sustainable integration, particularly as POC screening introduces additional demands on workflows, infrastructure, human resources, supply chains, and surveillance systems that affect feasibility and equity in practice.(20,21)

Few studies have examined implementation determinants of antenatal STI POC screening using implementation science frameworks. To address this gap, we conducted a mixed-methods process evaluation embedded within the Philani Ndiphile trial in the Eastern Cape, South Africa.(7,22) Guided by the Reach, Effectiveness, Adoption, Implementation, and Maintenance (RE-AIM) framework,(23) with the Consolidated Framework for Implementation Research informing conceptual analysis,(24) we assessed how POC STI screening functioned in a research setting and identified factors shaping implementation in government clinics from the perspectives of pregnant women, healthcare workers, and research staff.

## Methods

### Study design and setting

The Philani Ndiphile trial was a three-arm randomized controlled trial conducted between March 2021 and February 2025 across four public clinics in Buffalo City Municipality, Eastern Cape, South Africa.(22) It evaluated three antenatal STI screening strategies: baseline POC screening with a 21-day test-of-cure (Arm 1), baseline and 30–34 week POC screening (Arm 2), and standard syndromic management (Arm 3). The trial assessed impacts on birth outcomes, STI prevalence at delivery, and cost-effectiveness. We conducted a mixed-methods process evaluation embedded within the trial, guided by RE-AIM and informed by CFIR, using in-depth interviews and focus group discussions with pregnant women, healthcare workers, and research staff to identify contextual determinants of implementation.

#### Study Setting

Buffalo City Municipality (BCM) is a metropolitan area in South Africa’s Eastern Cape Province, with an estimated population of 893,157. It is a socioeconomically disadvantaged municipality with urban, peri-urban, and rural communities and a relatively young population.(25) The Eastern Cape has a high HIV burden, with an adult prevalence of 15.3% in 2017.(26) As of 2022, HIV prevalence among pregnant women was 36.7% in BCM, compared with 32.9% provincially and 27.5% nationally, while maternal syphilis prevalence was 3.0% in BCM, compared with 3.6% in the Eastern Cape and 2.6% nationally.(27)

The Philani Ndiphile trial was conducted across four government clinics in BCM: an urban community health centre (CHC) (Facility A), a high-volume urban primary healthcare centre (PHC) serving a peri-urban population (Facility B), a rural PHC serving vulnerable communities (Facility C), and a high-volume urban CHC serving a large peri-urban catchment (Facility D). (Figure 1) Images of the study sites have been published elsewhere.(29) In South Africa, PHC clinics provide first-contact preventive and curative services, while CHCs offer similar services with 24/7 availability and typically serve larger populations.(28)

**Figure 1.**
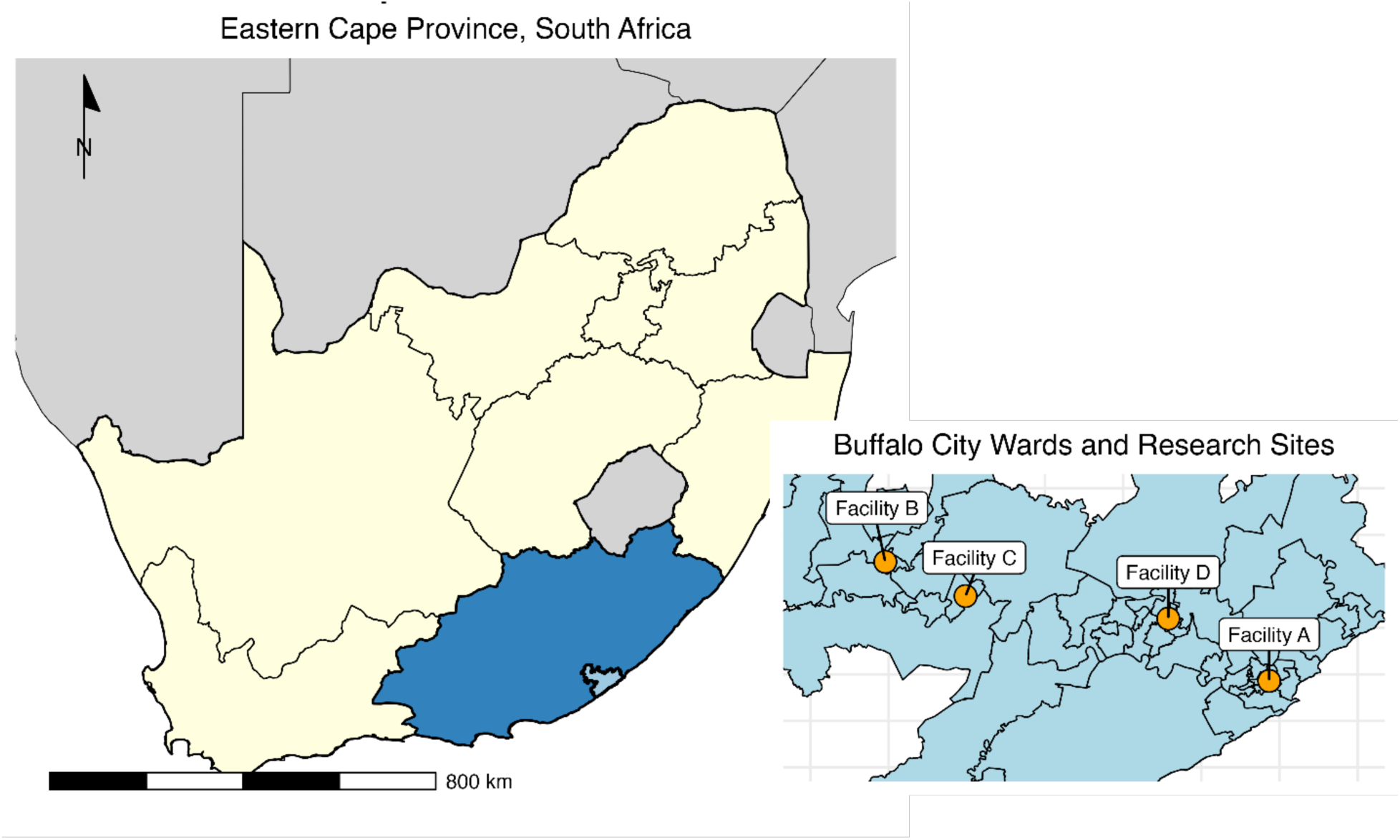
Philani Ndiphile Trial Research Sites, Buffalo City Municipality, Eastern Cape South Africa.

#### Study Population

The Philani Ndiphile trial enrolled consenting pregnant women attending their first ANC visit aged ≥18 years, <27 weeks’ gestation by ultrasound, and intending to deliver at a maternity obstetric unit within the municipality.(22)

The qualitative component included three stakeholder groups: pregnant women enrolled in the trial who received POC STI screening at baseline (Intervention Arms 1 and 2), research staff involved in trial procedures, and government healthcare workers and nurse managers involved in routine ANC service delivery at participating facilities. IDIs were conducted with pregnant women and FGDs with research staff and healthcare workers, with sample size guided by thematic saturation.

#### Process Evaluation and Theoretical Framework

This process evaluation was guided by the RE-AIM framework,(23) focusing on reach, adoption, implementation, and maintenance domains, with effectiveness outcomes reported elsewhere.(22) Quantitative process indicators included uptake of STI screening, treatment coverage among screen-positive women, same-day treatment coverage, and attendance at follow-up visits. These indicators were first analysed descriptively to characterise performance and to identify variation by facility.

CFIR(5) was used to identify contextual determinants underlying variation across RE-AIM domains. CFIR complemented RE-AIM by providing a detailed understanding of how contextual, facility, intervention, and individual factors influenced implementation. This integrated approach linked quantitative outcomes with qualitative insights to identify barriers and facilitators, compare across facilities, and assess requirements for sustained integration into government clinics. Process evaluation questions and measures are summarised in Supplementary File 1.

## Data Collection

### Quantitative data

Data were collected using REDCap,(25) with trained staff administering standardised questionnaires on sociodemographic, clinical, sexual health, obstetric, and process indicators. Research nurses conducted physical examinations and collected vaginal swabs from women enrolled in the intervention arms for screening of *C. trachomatis, N. gonorrhoeae*, and *T. vaginalis* using near point-of-care, electricity-dependent nucleic acid amplification test (GeneXpert CT/NG and TV assays; 90-minute turnaround time; Cepheid, Sunnyvale, California). Screen-positive women received treatment per South African guidelines,(9) while those in Arm 3 received standard-of-care syndromic management.(30) Notification slips were provided to women testing positive. All participants received routine ANC services,(31) including HIV and syphilis screening with confirmatory testing for those with positive screening results and immediate treatment.(32) Questions on intention to wait for STI screening results were added in September 2022, resulting in missing data for earlier enrolees.

### Qualitative data

Qualitative data were collected between September 2024 and April 2025. Pregnant women were purposively selected from a stratified random sample of trial participants and interviewed in isiXhosa or English at postnatal visits following informed consent. One FGD was conducted with research staff and four with government healthcare workers using purposive and convenience sampling. Interviews (∼20 minutes) and FGDs (∼1 hour) were conducted by trained female interviewers with no prior relationship to participants, using semi-structured guides informed by RE-AIM and CFIR [Supplemental File 2]. Interviews explored women’s experiences of STI screening, while FGDs examined facility workflows, resources, and implementation. Field notes documented contextual factors and group dynamics.

## Data Analysis

Quantitative analysis was conducted using R Studio Version 4.4.3 (2025-02-28). Process metrics were summarised descriptively. Differences in proportions were compared using Chi square or Fishers Exact tests, with a significance level of 0.05. Participant characteristics and process metrics were stratified by site to evaluate differences in populations and outcomes.

Interviews and FGDs were audio-recorded, transcribed, and translated, with accuracy checks by interviewers. Data were analysed using directed content analysis informed by CFIR, with two coders combining deductive and inductive coding in NVivo with a shared codebook. Coded themes were refined iteratively and examined across RE-AIM domains, with findings integrated using a weaving approach.

This study is reported in accordance with the Good Reporting of A Mixed Methods Study (GRAMMS)(33) and Consolidated Criteria for Reporting Qualitative Research (COREQ)(34) [Supplemental files 3 and 4].

## Ethics

Ethical approval was obtained from the University of Cape Town Faculty of Health Sciences Research Ethics Committee (ref. 676/2019). All Philani Ndiphile participants provided written informed consent in their preferred language (isiXhosa or English), with separate consent obtained for IDIs. All FGD participants provided individual informed consent. Participants were informed that participation was voluntary and would not affect access to care, study services, or employment. IDI participants received a R100 voucher; FGD participants were not reimbursed. All qualitative participants were informed of the goals of the research during the consenting process. A positionality and reflexivity statement can be found in Supplemental File 5.

## Patient and public involvement in research

Patients were not involved in study design or outcome development. Their perspectives informed findings through qualitative interviews on acceptability and experiences of POC STI screening. Participants contributed during study conduct but did not assess intervention burden. Results will be shared with participating facilities and communities through targeted dissemination.

## Results

### Participant characteristics

Overall, 2,247 women were enrolled in the Philani Ndiphile trial. Median age at enrolment was 28 years (IQR 24-33), median gestational age was 13 weeks (IQR 8-18), 41.7% were married or cohabitating with their partner, 43% were formally or self-employed, 29.1% were living with HIV, 3.05% had a positive syphilis rapid test result, and prevalence of any *C. trachomatis, N. gonorrhoeae*, and/or *T. vaginalis* among those in Arms 1 and 2 was 25.4% (379/1492) (Table1).

Twenty women participated in IDIs, median age was 30 years (IQR: 23.5–31.25) and median gestational age was 11 weeks (IQR: 8–16). Half of participants were recruited from Facility A (urban) (n = 10, 50%), while 40% (n = 8) were from peri-urban facilities (B and D) and 10% (n= 2) were from Facility C (rural). Participants were similar across trial arms (Arm 1 test-of-cure: 45%; Arm 2 repeat screen: 55%), baseline STI screening result waiting behaviour (waited: 55%; did not wait: 45%), and baseline STI positivity (any STI positive: 50%; negative: 50%). A total of 21 individuals participated in five FGDs, including one group with research staff (n = 7) and four groups with government healthcare workers (n = 14). Most FGD participants were female (n = 17, 81%) and professional nurses (n = 11, 52.3%). Among government healthcare workers, most were from Facilities A or B (10/14, 71.4%) (Supplemental File 6).

### Quantitative Process Outcomes

#### Uptake of screening

The proportion of eligible participants who consented to participate in the trial was 99% (2,396/2,419). Reasons for non-participation included fear of trial procedures(n = 6), lack of time (n = 2), or other unspecified reasons (n = 14).

#### Treatment coverage

The proportion of STI screen-positive women who received treatment was 95.2% at baseline and 93.5% at repeat screening (Figure 2). Treatment coverage across facilities ranged from 89.1% at Facility D (peri-urban CHC) to 100% at Facility C (rural PHC) at baseline (p = 0.011), and from 85.7% at Facility D (peri-urban CHC) to 100% at Facilities B (peri-urban PHC) and C (rural PHC) at repeat screening (p = 0.32) (Table 2).

**Figure 2.**
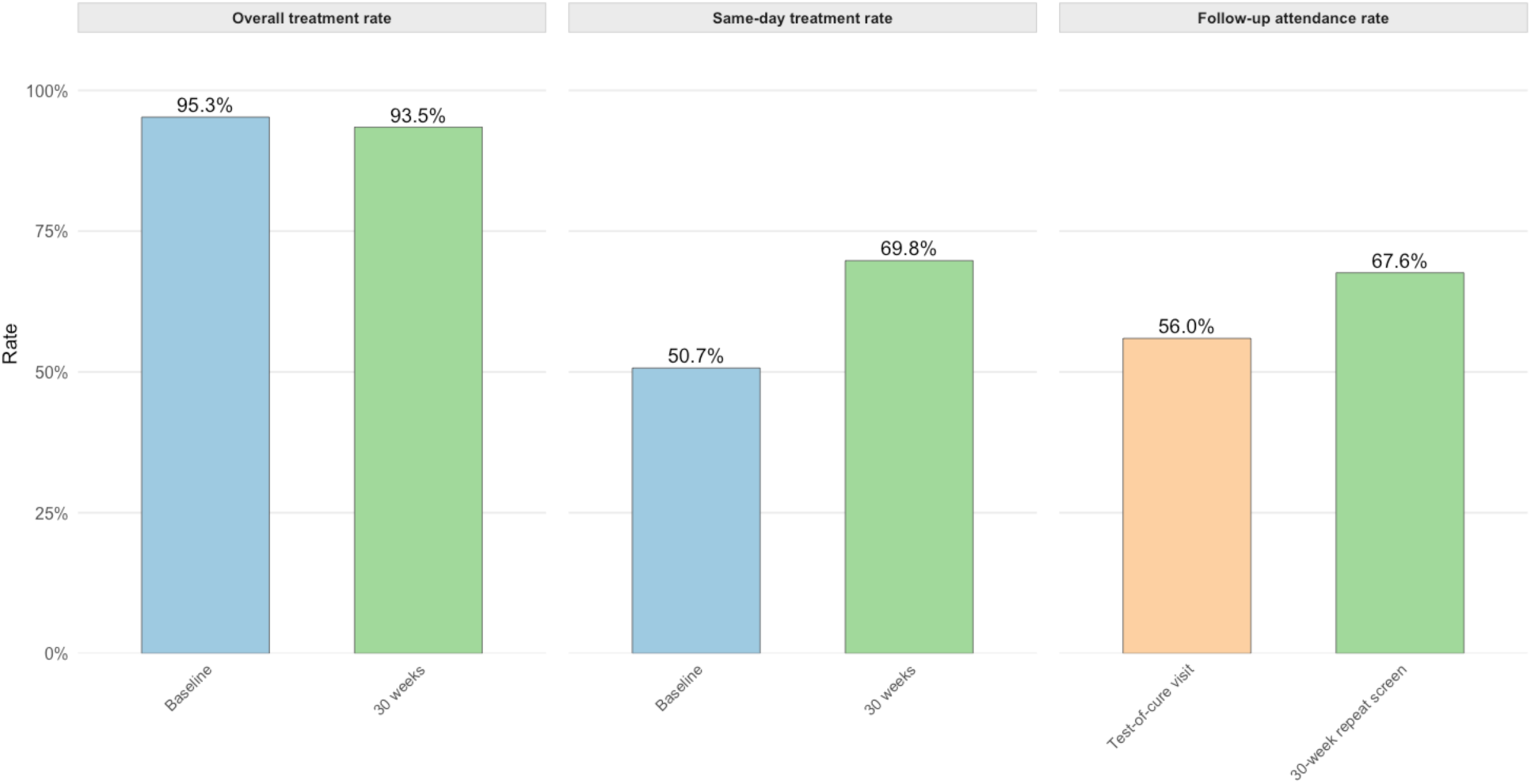
Overall treatment rate, same-day treatment rate, and follow-up attendance rates among participants enrolled in the Philani Ndiphile Trial.

#### Same-day treatment coverage

The proportion of POC STI screen-positive women who received treatment on the same-day as their POC test was 50.7% at baseline and 69.8% at repeat screening (Figure 2). Facility-level coverage ranged from 10% at Facility D (peri-urban CHC) to 90.7% at Facility B (peri-urban PHC) (p < 0.01) at baseline and increased across all facilities at the repeat visit, though remained lower at Facility D (25%) (p < 0.01) (Table 2).

#### Follow-up attendance rates

The proportion of all expected women in Arm 1 who attended their 21-day test-of-cure visit (n = 108/193, 56%) was lower than the proportion of expected women in Arm 2 who attended the 30-34 week repeat screening visit (n = 499/738, 68%) (p < 0.01). Follow-up attendance at the routine 30-34 week ANC visit was similar among all trial participants (n = 1509/2247, 67%) compared to only those in the repeat screening arm (n = 499/738, 68%) (Figure 2), and varied by site, ranging from 58.8% at Facility D (peri-urban CHC) to 75.7% at Facility B (peri-urban PHC) (p < 0.01) (Table 2).

### Mixed-methods findings

To contextualise quantitative findings and explain observed differences, we present qualitative insights from pregnant women, research staff, and healthcare workers, highlighting determinants of POC STI screening implementation in ANC. Key findings mapped to CFIR domains within the RE-AIM framework are summarised in Figure 3.

**Figure 3:**
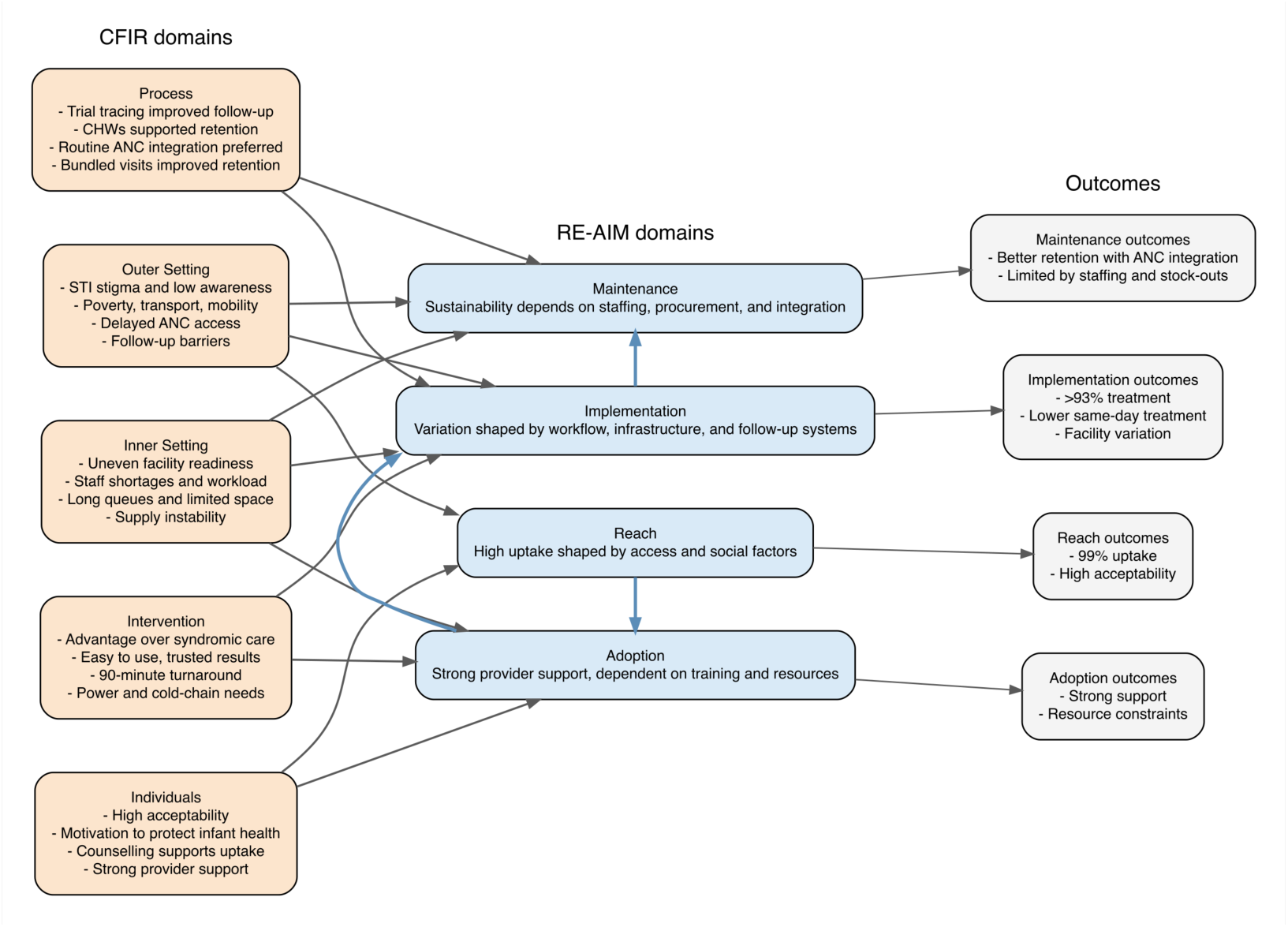
Conceptual framework of determinants influencing implementation of antenatal point-of-care STI screening across RE-AIM domains.

### Reach

Reach assessed access to and engagement with STI screening among pregnant women, the representativeness of those reached, and the acceptability of testing among eligible participants.

#### Acceptance among pregnant women

Pregnant women expressed positive attitudes toward POC STI screening and treatment, motivated by protecting their own health and that of their baby, and valuing early detection and prevention: *“…if I have a disease, it could be discovered early.”* (Participant 2, Facility A – urban CHC)

Although some women reported initial anxiety, supportive, person-centred counselling and clear communication from research nurses facilitated uptake, engagement, and increased knowledge of STI testing:

> *“I was nervous about the results, but then they assured me… if they find anything it can be cured.”* (Participant 3, Facility C – rural)

#### Sociocultural and Structural Barriers to Access

Research staff and government providers identified limited community awareness and stigma around STIs and low STI risk awareness as key barriers to reaching women. Research staff emphasised the need for outreach to improve awareness and generate demand, including efforts to *“go to the communities and speak to people”* (Research Field Officer 2).

Poverty, low health literacy, and delayed ANC booking were perceived as barriers to uptake of ANC more broadly, with some providers perceiving women in their communities to have limited awareness of the importance of early care:

> *“We are dealing with a community with a high rate of unemployment and low education level… they are pregnant, but there is no insight that they must come early”* (Nurse Manager, Facility D – peri-urban CHC).

Socioeconomic factors affected ANC engagement: 68.3% lacked private transport, median travel time was 20 minutes, and 12.7% at the rural facility (C) travelled over an hour. Clinic visits also incurred costs, with 16.6% reporting income loss and 75.8% transport expenses. (Table 1)

**Table 1.** Baseline characteristics of pregnant women enrolled in the Philani Ndiphile Trial in Eastern Cape, South Africa (N=2247)

| Participant characteristic | Facility |  |  |  |  |
| --- | --- | --- | --- | --- | --- |
|  | Total<sup>1</sup><br>N = 2247<sup>1</sup> | A (urban)<br>N = 667<sup>1</sup> | B (peri-urban)<br>N = 691<sup>1</sup> | C (rural)<br>N = 251<sup>1</sup> | D (peri-urban)<br>N = 638<sup>1</sup> |
| **Age (years)** | 28 (24, 33) | 28 (24, 33) | 28 (23, 33) | 28 (23, 34) | 28 (23, 33) |
| **Gestational age (weeks)** | 13.0 (8.0, 18.0) | 13.0 (8.0, 18.0) | 13.0 (9.0, 18.0) | 14.0 (8.0, 18.0) | 14.0 (8.0, 19.0) |
| **Living with HIV** | 655 (29.1%) | 219 (32.8%) | 223 (32.3%) | 98 (39.0%) | 115 (18.0%) |
| **On ART at first ANC†** | 535 (81.7%) | 175 (79.9%) | 192 (86.1%) | 79 (80.6%) | 89 (77.4%) |
| **Positive for any STI\*** | 379 (25.4%) | 113 (25.5%) | 112 (24.3%) | 53 (32.9%) | 101 (23.7%) |
| **Highest education level** |  |  |  |  |  |
| Gr 10-12 | 1,798 (80.1%) | 541 (81.2%) | 526 (76.2%) | 210 (83.7%) | 521 (81.7%) |
| Higher education | 289 (12.9%) | 71 (10.7%) | 133 (19.3%) | 14 (5.6%) | 71 (11.1%) |
| Less than gr 10 | 158 (7.0%) | 54 (8.1%) | 31 (4.5%) | 27 (10.8%) | 46 (7.2%) |
| *Missing* | *2* | *1* | *1* | *0* | *0* |
| **Married or cohabitating with partner** | 938 (41.7%) | 317 (47.5%) | 245 (35.5%) | 80 (31.9%) | 296 (46.4%) |
| **Currently employed (including self-employed)** | 966 (43.0%) | 281 (42.1%) | 337 (48.8%) | 58 (23.1%) | 290 (45.5%) |
| **Personal income§** |  |  |  |  |  |
| < 1000 ZAR per month | 102 (10.6%) | 27 (9.6%) | 26 (7.7%) | 3 (5.2%) | 46 (15.9%) |
| >10 000 ZAR per month | 51 (5.3%) | 13 (4.6%) | 22 (6.5%) | 0 (0.0%) | 16 (5.5%) |
| 1001–5000 ZAR per month | 631 (65.3%) | 186 (66.2%) | 218 (64.7%) | 48 (82.8%) | 179 (61.7%) |
| 5001–10 000 ZAR per month | 174 (18.0%) | 55 (19.6%) | 68 (20.2%) | 6 (10.3%) | 45 (15.5%) |
| None | 8 (0.8%) | 0 (0.0%) | 3 (0.9%) | 1 (1.7%) | 4 (1.4%) |
| **Needed to borrow money to fund pregnancy** | 281 (12.5%) | 58 (8.7%) | 126 (18.2%) | 63 (25.1%) | 34 (5.3%) |
| **Has access to working private transport (vehicle or bicycle)** | 712 (31.7%) | 196 (29.4%) | 292 (42.3%) | 53 (21.1%) | 171 (26.8%) |
|  | Total<sup>1</sup><br>N = 2247<sup>1</sup> | A (urban)<br>N = 667<sup>1</sup> | B (peri-urban)<br>N = 691<sup>1</sup> | C (rural)<br>N = 251<sup>1</sup> | D (peri-urban)<br>N = 638<sup>1</sup> |
| **Transport cost to clinic** |  |  |  |  |  |
| None | 704 (31.3%) | 225 (33.7%) | 205 (29.7%) | 158 (62.9%) | 116 (18.2%) |
| ≤50 ZAR | 1,202 (53.5%) | 355 (53.2%) | 398 (57.6%) | 90 (35.9%) | 359 (56.3%) |
| 51–150 ZAR | 307 (13.7%) | 83 (12.4%) | 79 (11.4%) | 2 (0.8%) | 143 (22.4%) |
| >150 ZAR | 34 (1.5%) | 4 (0.6%) | 9 (1.3%) | 1 (0.4%) | 20 (3.1%) |
| **Lost income because of coming to the clinic§** | 160 (16.6%) | 51 (18.1%) | 89 (26.4%) | 12 (20.7%) | 8 (2.8%) |
| **Amount of income lost due to clinic visit** |  |  |  |  |  |
| ≤100 ZAR | 42 (26.3%) | 11 (21.6%) | 26 (29.2%) | 1 (8.3%) | 4 (50.0%) |
| 101–400 ZAR | 100 (62.5%) | 32 (62.7%) | 53 (59.6%) | 11 (91.7%) | 4 (50.0%) |
| 401–800 ZAR | 14 (8.8%) | 5 (9.8%) | 9 (10.1%) | 0 (0.0%) | 0 (0.0%) |
| >800 ZAR | 4 (2.5%) | 3 (5.9%) | 1 (1.1%) | 0 (0.0%) | 0 (0.0%) |
| **Travel time to clinic** |  |  |  |  |  |
| ≤10 | 377 (16.8%) | 136 (20.4%) | 113 (16.4%) | 25 (10.0%) | 103 (16.1%) |
| 11-30 | 1,476 (65.7%) | 426 (63.9%) | 453 (65.6%) | 128 (51.0%) | 469 (73.5%) |
| 31-60 | 325 (14.5%) | 93 (13.9%) | 108 (15.6%) | 66 (26.3%) | 58 (9.1%) |
| >60 | 69 (3.1%) | 12 (1.8%) | 17 (2.5%) | 32 (12.7%) | 8 (1.3%) |
| **Time normally spent in clinic queue** |  |  |  |  |  |
| <30 | 847 (37.7%) | 414 (62.1%) | 296 (42.8%) | 106 (42.2%) | 31 (4.9%) |
| 31–60 | 442 (19.7%) | 121 (18.1%) | 222 (32.1%) | 41 (16.3%) | 58 (9.1%) |
| >60 | 958 (42.6%) | 132 (19.8%) | 173 (25.0%) | 104 (41.4%) | 549 (86.1%) |
| **Number of sexual partners (past 12 months)** |  |  |  |  |  |
| More than one | 264 (11.7%) | 74 (11.1%) | 82 (11.9%) | 33 (13.1%) | 75 (11.8%) |
| One | 1,983 (88.3%) | 593 (88.9%) | 609 (88.1%) | 218 (86.9%) | 563 (88.2%) |
|  | Total<sup>1</sup><br>N = 2247<sup>1</sup> | A (urban)<br>N = 667<sup>1</sup> | B (peri-urban)<br>N = 691<sup>1</sup> | C (rural)<br>N = 251<sup>1</sup> | D (peri-urban)<br>N = 638<sup>1</sup> |
| **Suspect partner has other sexual partners±** |  |  |  |  |  |
| No | 1,177 (55.2%) | 395 (60.7%) | 328 (50.2%) | 111 (47.2%) | 343 (58.0%) |
| Unsure | 344 (16.1%) | 70 (10.8%) | 109 (16.7%) | 53 (22.6%) | 112 (19.0%) |
| Yes | 610 (28.6%) | 186 (28.6%) | 217 (33.2%) | 71 (30.2%) | 136 (23.0%) |
| **Would disclose STI results to partner** |  |  |  |  |  |
| Yes, to steady partner | 2,196 (97.7%) | 656 (98.4%) | 679 (98.3%) | 247 (98.4%) | 614 (96.2%) |
| Yes, to casual partner(s) | 11 (0.5%) | 0 (0.0%) | 0 (0.0%) | 0 (0.0%) | 11 (1.7%) |
| Yes, to all steady and casual partner(s) | 1 (0.0%) | 0 (0.0%) | 0 (0.0%) | 0 (0.0%) | 1 (0.2%) |
| No | 39 (1.7%) | 11 (1.6%) | 12 (1.7%) | 4 (1.6%) | 12 (1.9%) |
| **Used a condom at last sex** | 226 (10.1%) | 84 (12.6%) | 73 (10.6%) | 27 (10.8%) | 42 (6.6%) |
| **Intending to wait for STI test results‡** | 796 (59.0%) | 245 (58.8%) | 333 (93.0%) | 204 (92.7%) | 14 (4.0%) |
| *Missing* | *898* | *250* | *333* | *31* | *284* |
| **Reason for not intending to wait for results‡** |  |  |  |  |  |
| Have to get to work | 146 (26.4%) | 35 (20.3%) | 9 (36.0%) | 3 (18.8%) | 99 (29.1%) |
| Hungry | 112 (20.3%) | 38 (22.1%) | 0 (0.0%) | 5 (31.3%) | 69 (20.3%) |
| Have to get back to my kids/family | 81 (14.6%) | 29 (16.9%) | 8 (32.0%) | 6 (37.5%) | 38 (11.2%) |
| Not feeling well | 62 (11.2%) | 16 (9.3%) | 0 (0.0%) | 0 (0.0%) | 46 (13.5%) |
| Want to go to the shop | 51 (9.2%) | 12 (7.0%) | 1 (4.0%) | 0 (0.0%) | 38 (11.2%) |
| Transport availability | 4 (0.7%) | 1 (0.6%) | 1 (4.0%) | 0 (0.0%) | 2 (0.6%) |
| Boring | 2 (0.4%) | 1 (0.6%) | 0 (0.0%) | 0 (0.0%) | 1 (0.3%) |
| No space to wait | 2 (0.4%) | 2 (1.2%) | 0 (0.0%) | 0 (0.0%) | 0 (0.0%) |
| Other | 93 (16.8%) | 38 (22.1%) | 6 (24.0%) | 2 (12.5%) | 47 (13.8%) |
|  | Total<sup>1</sup><br>N = 2247<sup>1</sup> | A (urban)<br>N = 667<sup>1</sup> | B (peri-urban)<br>N = 691<sup>1</sup> | C (rural)<br>N = 251<sup>1</sup> | D (peri-urban)<br>N = 638<sup>1</sup> |
| *Missing* | *1,694* | *495* | *666* | *235* | *298* |

<sup>1</sup>Median (Q1, Q3); n (%)
\* Only among participants in Intervention Arms 1 and 2 who received aetiological testing at the baseline visit. Any STI includes *Chlamydia trachomatis*, *Neisseria gonorrhoea*, or *Trichomonas vaginalis*
§ Only asked among those who reporting being employed
± Only asked among those who reported having a steady partner
† Among participants living with HIV
‡ Question added to study questionnaire on 13/09/2022

**Table 2.** Philani Ndiphile Study Process Metrics by Facility.

|  | Facility |  |  |  |  | p-value |
| --- | --- | --- | --- | --- | --- | --- |
|  | Total<sup>1</sup> | A<sup>1</sup> | B<sup>1</sup> | C<sup>1</sup> | D<sup>1</sup> |  |
| **Positive for any STI at baseline (N)** | **379** | **113** | **112** | **53** | **101** |  |
| Received treatment at any timepoint | 361<br>(95.3%) | 109<br>(97.3%) | 109<br>(96.4%) | 53<br>(100%) | 90<br>(89.1%) | 0.011 |
| Received same-day treatment | 183<br>(50.7%) | 29<br>(26.6%) | 99<br>(90.8%) | 46<br>(86.8%) | 9<br>(10.1%) | < 0.01 |
| **Positive for any STI at 30-week visit (N)** | **46** | **9** | **15** | **8** | **14** |  |
| Received treatment at any timepoint | 43<br>(93.5%) | 8<br>(89.9%) | 15<br>(100%) | 8<br>(100%) | 12<br>(85.7%) | 0.32 |
| Received same-day treatment | 30<br>(69.8%) | 4 (50%) | 15<br>(100%) | 8<br>(100%) | 3 (25%) | < 0.01 |
| **Expected at routine 30-week ANC visit (all study arms) (N)** | **2247** | **667** | **691** | **251** | **638** |  |
| Attended routine 30-week visit | 1509<br>(67.2%) | 424<br>(63.6%) | 523<br>(75.7%) | 187<br>(74.5%) | 375<br>(58.8%) | < 0.01 |

<sup>1</sup>n (%)

## Adoption

Adoption examined the willingness and readiness of healthcare workers and facilities to implement aetiological STI screening within routine ANC.

### Provider willingness

Healthcare workers expressed strong willingness to adopt POC STI screening, driven by perceived improvements in clinical care and professional identity. Government providers viewed aetiological screening as critical for improving the health of pregnant women in their care, noting that study staff often caught infections when syndromic management failed:

> *“Most of the time, the client would not know she has a problem. Then when they go for screening, [study staff] will pick up and inform us [the client] has an STI and we need to get treatment ready.”* – (Professional Nurse, Facility B – peri-urban PHC)

The use of new technology and skill development was a strong motivator for healthcare workers, especially among younger nurses, who were perceived as eager to engage with new technologies: *“… they would be up in arms getting to know this tool.”* (Professional Nurse, Facility D – peri-urban HC)

### Relative advantage and usability

Research and government providers described aetiological screening as superior to syndromic management, as it identified otherwise undetected infections with implications for maternal and neonatal health:

> *“There is absolutely nothing abnormal about this participant… yet she tests positive for one, two, or even all three STIs. To think this woman would have given birth without knowing—and possibly infected her partner—is really quite phenomenal”* (Research Nurse).

High usability supported provider acceptance, with clear procedures, straightforward results, and few errors with the GeneXpert platform linked to provider acceptance: *“Simple is best.”* – (Professional Nurse, Facility A – urban CHC)

Healthcare workers and pregnant women expressed confidence in provider-collected vaginal swabs, viewing them as more reliable despite some discomfort: *“it is better when the nurse does it because I would think I am doing the right thing, but I am not”* (Participant 1, Facility A – urban CHC). While some healthcare workers felt swab collection could be integrated into routine antenatal exams, others questioned its feasibility at scale given high patient volumes and limited consultation time.

### Organisational support

Government healthcare workers expressed strong willingness to transition from syndromic management to aetiological STI screening, provided adequate resources, training, and institutional support were in place. While aligned with ANC goals, providers in high-volume clinics anticipated resistance due to workload pressures and noted that scale-up would depend on staffing capacity and workflow integration.

> *“We are a very high-volume facility, by the end of the day we are exhausted… there might be slight resistance due to the volume of patients we see daily.”* (Professional Nurse, Facility B – peri-urban PHC)

Adequate training was considered essential, with support for modular or rotational models and broad cadre training to enable task-shifting without disrupting services, with one government healthcare worker noting, *“…eventually all of us should be trained.”* (Professional Nurse, Facility D – peri-urban CHC)

### Implementation

Implementation examined how POC STI screening was delivered during the trial, focusing on feasibility, fidelity, and consistency of delivery across sites, as well as adaptations and systems needed for integration within government facilities.

#### Resources and infrastructure

Infrastructure, equipment, and cold-chain requirements influenced perceived feasibility. Although some providers felt the compact GeneXpert platform could help address space constraints, others, particularly in high-volume clinics, described facilities as physically *“restricted”* and unable to accommodate additional equipment or the laptops needed for test processing and results.

Intermittent electricity reduced confidence in the platform’s continuous power requirement. While most facilities were affected mainly during national power outages, the rural facility reported more frequent disruptions, noting that *“sometimes the plugs will just not work”* (Professional Nurse, Facility C – rural). During the trial, research nurses adapted testing schedules around power cuts, which they reported led to delayed processing times. Cold-chain requirements also raised feasibility concerns, as government facilities largely relied on cooler boxes, which providers felt may be insufficient at higher testing volumes or for stricter storage conditions.

#### Turnaround time

Research staff identified that the 90-minute GeneXpert processing time limited same-day treatment, noting that women who did not wait often received delayed treatment or were lost to follow-up:

> *“You’d treat them after a week or so or not treat them at all because they don’t come back” (*Research Nurse, Facility D – peri-urban CHC).

Providers at high-volume sites described the processing time as incompatible with busy workflows and limited waiting space; however some felt targeted counselling could increase willingness to wait, particularly when framed around foetal health benefits: *“If pregnant women know the benefits… they will wait”* (Professional Nurse, Facility A – urban CHC). However, those

Some women were willing to wait to avoid additional visits and receive immediate treatment, *“… I am not going to come here again on another day.”* (Participant 5, Facility A – urban CHC) However, competing demands such as work, childcare, hunger, and long clinic waits limited this. Overall, 40.9% did not intend to wait at baseline, most commonly due to employment commitments (24.4%) and hunger (20.3%), while lack of waiting space (0.4%) and boredom (0.4%) were rarely cited. (Table 1)

#### Follow-up constraints

Loss to follow-up was a challenge during the trial. Study staff reported difficulties tracing women due to phone theft in urban areas and long distances, mobility, and poor network coverage in peri-urban and rural settings. Research staff employed strategies to ensure treatment completion and retention, including obtaining several alternative phone numbers and tracking women’s clinic follow-up appointments, however government providers felt human resources were insufficient to sustain intensive tracing, particularly in large, mobile catchment areas:

> *“…the patient is not coming back… they jump from space to space”* (Professional Nurse, Facility D – peri-urban CHC).

Several facilities used community health workers (CHW) to support treatment outreach and follow-up. Government providers at the rural and urban sites viewed this as a natural extension of existing community support, with one noting, *“We rely a lot on [CHW] to help reach women.”* (Professional Nurse, Facility C – rural). However, facilities serving large catchment populations reported that CHWs were already overburdened with tracing activities and that adding STI follow-up would be unsustainable.

#### Facility readiness

Implementation varied across sites due to differences in facility readiness, infrastructure, patient volume, and catchment characteristics. Research staff reported that collaboration with facility providers, clinic capacity, and local population factors influenced delivery and follow-up. At high-volume facilities, healthcare workers cited long queues and constrained workflows as barriers to engagement and follow-up.

> *“… it’s a challenge for them just to wait for the nurse. One hour of queuing and the client is already panicking.”* (Professional Nurse, Facility A – urban CHC)

This was consistent with trial data, which showed women waited a median of 60 minutes (IQR: 30–119) to see a provider; with 42.6% of women waiting more than one hour, increasing to 86.1% at Facility D (peri-urban CHC) (Table 1).

### Maintenance

Maintenance examined the conditions required for sustained delivery and effectiveness of POC STI screening beyond the research setting.

#### Resource sustainability

During the trial, GeneXpert platforms were study-supplied. Government providers questioned the feasibility of sustaining these platforms in public clinics. Concerns centred on unclear accountability for maintenance and procurement within the constrained public health system. Frequent stock-outs were cited as a major barrier to consistent POC screening, with facilities often reverting to laboratory-based testing resulting in delaying treatment initiation: “*we used to have that rapid [syphilis] test… now we need to draw blood and wait for the results*” (Professional Nurse, Facility C – rural).

Human resource shortages were pervasive across facilities. During the trial, research staff conducted all study procedures and provided routine ANC services to participants.

Government providers reported that absorbing POC STI screening tasks without additional staff would be unsustainable given existing workloads:

> *“If we implement this tool, it will add extra work on the shoulders of the employees… We would need an extra hand for the for the program to be sustainable. If we get an extra job, in addition to the jobs that we are doing right now, I assure you, it cannot be sustainable”* (Nurse Manager, Facility D – peri-urban CHC).

#### Reinfection risk

Reinfection emerged as a key threat to sustained effectiveness. Providers reported that partner notification and treatment were constrained by gender norms, STI-related stigma, and low male involvement in ANC, placing the burden on pregnant women. Male partners rarely attended ANC and were often unwilling to wait for results or accept treatment due to work commitments. While 98% of trial participants were willing to disclose results to a stable partner, fewer than 1% would disclose to a casual partner and 2% would not disclose at all. (Table 1)

Providers also reported limited ability to negotiate condom use, particularly among economically dependent women; consistent with this, only 10% reported condom use at last sex, increasing the risk of reinfection and reducing sustained impact. (Table 1)

> *“The partners don’t want to use [condoms], and she also doesn’t say no to this boyfriend because they depend on him. Maybe she’s not working and then she doesn’t want to lose this boyfriend”* (Professional Nurse, Facility C – rural).

#### Integration

Healthcare workers and pregnant women emphasised that sustainability depended on integrating STI screening into existing service touchpoints. Bundling testing within routine ANC visits was preferred over standalone appointments, which were viewed as burdensome:

> *“[Testing should be conducted] during antenatal care visits, because I’m coming here anyway. I don’t like doing frequent visits”* (Participant 11, Facility B – peri-urban PHC).

Consistent with this, attendance was higher for repeat screening integrated into the 30–34-week ANC visit than for non-routine test-of-cure visits (Figure 2). Women also supported integrating STI screening into other routine services, including family planning, well-baby, and ART visits: *“I will not always be pregnant, but I always have to come for family planning”* (Participant 8, Facility B – peri-urban PHC).

#### Screening Frequency

Early universal screening was viewed as important for preventing missed infections, though healthcare workers in high-volume settings questioned feasibility of universal screening at scale and preferred risk-based screening: *“If we were to screen everyone, that would not be feasible.”* (Professional Nurse, Facility B – peri-urban PHC).

Healthcare workers and women also viewed repeat screening later in pregnancy as essential for preventing reinfection and protecting maternal and infant health:

> *“I would like to be tested at every visit… To be safe for my baby”* (Participant 9, Facility D – peri-urban CHC)

Women were willing to return for follow-up screening, motivated by protecting their baby’s health. Despite no reported barriers to returning during IDIs, retention during the trial remained limited, with 67% attending the 30–34-week visit and ∼25% attrition even at the best-performing facility (B – urban CHC). (Table 2).

## Discussion

This mixed-methods process evaluation examined how antenatal STI screening using a near point-of-care, electricity-dependent nucleic acid amplification platform functioned in a research setting and identifies key factors for implementation in routine ANC. Our findings show that POC STI screening was highly acceptable and feasible. However, uptake, adoption, implementation, and sustainability in government clinics will depend on facility capacity, assay requirements, clinic workflows, and broader socioeconomic and structural factors shaping engagement across the care cascade.

We observed near-universal uptake of POC STI screening among eligible pregnant women within the context of a research study. While a small number cited fear of testing as a reason for non-participation, person-centred counselling reduced anxiety and facilitated engagement, consistent with prior evidence from South Africa.(35) However, broader geographic and socioeconomic barriers, including transport costs, lost income, and food insecurity, may limit access to antenatal STI screening. These structural constraints, which affect ANC attendance more broadly,(36) contribute to missed opportunities for preventing vertical transmission,(37) highlighting the need to address basic needs to ensure equitable reach and impact.

High treatment coverage at baseline and 30-week visits adds to evidence supporting the effectiveness of STI test-and-treat approaches among women engaged in care.(15,38) However, treatment gaps persisted at Facility D, with qualitative findings indicating that this peri-urban CHC serves a highly mobile population and faces constraints such as long queues and limited infrastructure. Similar challenges in antenatal syphilis programmes persist in South Africa,(37) and have been linked to facility-level challenges such as stock-outs, staffing shortages, and workflow limitations.(39–42) These findings highlight the risk of loss to follow-up when treatment is delayed and underscore the need for strengthened workflows and supply systems to support immediate care.

Despite high treatment coverage, same-day treatment, a key component of test-and-treat strategies, remained lower. The 90-minute turnaround time was a major barrier, consistent with evidence linking longer GeneXpert processing times to reduced same-day uptake.(43–48) However, high same-day treatment rates reported in similar settings using the same platform(13,38,49) suggest that turnaround time alone does not explain performance differences. We observed substantial facility-level variation; although providers cited space constraints, these were rarely linked to women’s willingness to wait, which was more strongly influenced by hunger and work commitments, and prior work from our team showed that waiting areas alone did not improve same-day treatment uptake.(29). These findings highlight the need for context-adapted delivery models alongside more rapid diagnostics. Emerging aetiological tests with shorter turnaround times, including lateral flow assays for N. gonorrhoeae, may help address operational constraints associated with longer processing times and electricity-dependent platforms.(50,51)

Although healthcare workers supported aetiological screening and recognised its advantages over syndromic management, readiness for scale-up depended on facility capacity to meet platform requirements. Evidence from Uganda suggests provider willingness alone is insufficient without assays adapted to local contexts and integrated into health systems.(52) While provider-collected swabs were widely accepted, their feasibility in high-volume settings may be limited. Self-collected vaginal swabs offer a viable alternative, with evidence showing high acceptability when combined with clear instruction,(53–55) comparable accuracy profiles,(56) and potential to reduce workforce burden while supporting decentralised delivery.(57–59) These findings highlight the need for simplified, context-appropriate diagnostic approaches and user-informed implementation strategies.

In our study, perceived feasibility and sustainability of POC STI screening depended on alignment with existing resources. Human resource constraints were identified as a key barrier, consistent with evidence from similar settings showing that workforce instability undermines protocol adherence, training continuity, quality of care, and waiting times for POC screening.(60–64) Healthcare workers supported cross-cadre training to facilitate task-shifting and improve implementation feasibility. Task-shifting can be cost-effective in low-resource settings,(65) with studies from South Africa and similar contexts showing that reallocating low-complexity tasks to lay staff reduces workforce pressures and allows nurses to focus on specialised care.(66–69) However, evidence remains largely limited to HIV services, highlighting the need for context-specific evaluations for STI screening.

Our findings suggest that POC STI screening should be integrated into existing clinical workflows, consistent with evidence that bundled HIV, syphilis, and hepatitis B screening is cost-effective(70) and improves test uptake.(71) Although evidence for other STIs is limited, data from Zimbabwe support the feasibility of integrating *C. trachomatis, N. gonorrhoeae*, and *T. vaginalis* screening within routine prevention of vertical transmission services.(38) Repeat screening was also acceptable when aligned with routine ANC visits, but attrition limited sustained impact, particularly in high-mobility setting. Similar retention gaps have been reported for repeat HIV and syphilis screening algorithms in South Africa,(37,64,72) reflecting persistent system-level barriers to longitudinal care. In our study, Facilities using CHWs achieved higher retention, consistent with evidence of their effectiveness in improving ANC attendance and adherence.(71,73) These findings highlight the importance of integrated, community-linked models for longitudinal POC STI screening, follow-up, and surveillance.

This study has several limitations. It was conducted in a single municipality and excluded women younger than 18 years and those presenting later to ANC, limiting generalisability. Interviews were mainly conducted at one high-performing urban facility, potentially underrepresenting rural and peri-urban experiences. Participants experienced only provider-collected swabs, so comparisons with self-collection were hypothetical. Responses may also be affected by social desirability and recall bias.

## Conclusion

Aetiological POC screening for curable STIs using near point-of-care, electricity-dependent nucleic acid amplification platforms was acceptable and feasible among pregnant women, achieving high uptake and treatment coverage in a research setting. However, sustainable scale-up in government clinics will depend on local adaptation, workflow integration, human resources, reliable supply chains, effective follow-up systems, and rapid diagnostic platforms aligned with routine ANC workflows. Without addressing persistent gaps in ANC access, facility readiness, and retention across the care cascade, the benefits of STI POC screening may be unevenly realised, further widening existing maternal and neonatal health disparities.

## Declarations

## Acknowledgements

We would like to acknowledge and thank the participants in our study and the Foundation for Professional Development staff in East London who worked on this study.

## Ethical considerations

Ethical approval was provided by the Institutional Review Boards at the University of Cape Town’s Faculty of Health Sciences Research Ethics Committee (UCT-HREC, reference number 676/2019).

## Consent to participate

All participants provided written informed consent in their preferred language (isiXhosa or English) prior to randomization.

## Consent for publication

Not applicable

## Declaration of confficting interest

The author(s) declared no potential conflicts of interest with respect to the research, authorship, and/or publication of this article

## Data availability

The data that support the findings of this study are available from the corresponding author upon reasonable request.

## Supplemental Files

**Supplemental File 1:** RE-AIM process evaluation questions, measures, and mapped CFIR findings

**Supplemental File 2:** Semi-Structured Interview Guides for interviews and focus group discussions

**Supplemental File 3:** COREQ Checklist

**Supplemental File 4:** GRAMMS Checklist

**Supplemental File 5:** Positionality and Reflexivity Statement

**Supplemental File 6:** Characteristics of Interview and Focus Group Participants

