## Supplemental File 1 for "Implementation of point-of-care screening for *Chlamydia trachomatis, Neisseria gonorrhoeae*, and *Trichomonas vaginalis* among pregnant women in South Africa: a mixed-methods process evaluation of the Philani Ndiphile trial"

**Supplemental File 1: Key findings mapped to CFIR and RE-AIM Frameworks**

| **Key Evaluation Questions** | **Quantitative Indicators** | **Quantitative measures** | **CFIR Domain(s)** | **Key Themes** | **Illustrative Quotations** |
| --- | --- | --- | --- | --- | --- |
| **Reach**  *The proportion and representativeness of individuals who are willing to participate in STI POCT* | | | | | |
| 1. Who was reached by STI POCT? 2. Which groups were less likely to attend follow-up or wait for results? | - Proportion of eligible pregnant women receiving STI POCT at baseline - Attendance at routine 30-week ANC visit and non-routine test-of-cure visit - Reach stratified by site, age, gestational age, and socioeconomic indicators (transport access, income loss) - Access-related barriers, including transportation and cost | - Women’s perceptions of access to ANC and STI POCT. - Woman’s perceptions of STI POCT - Community- and partner-level influences on engagement | Characteristics of pregnant women | - High acceptability of STI POCT - Motivation driven by fetal and maternal health protection - Initial anxiety mitigated by supportive counselling - Trust in providers facilitated acceptance - Increased STI knowledge and health literacy - Enhanced sense of agency and self-efficacy | - *“…Even though I was nervous at first, I did calm down after they talked to me.”* – (Participant 11) - *“I wanted to be safe… so that I give birth to a healthy child who is not infected”* – (Participant 6) |
|  |  |  | Outer Setting; | - Limited community awareness and stigma - Socioeconomic vulnerability shaping care-seeking behaviour - Geographic and transport barriers to ANC access | - *“We are dealing with a community with a high rate of unemployment and low education level… they are pregnant, but there is no insight that they must come early.”* – (Nurse Manager, Facility D) - *“If we could have more campaigns whereby it will be spoken among the community , so that that people can know they can come to us at the facility”* – (Research Nurse 2) |
| **Adoption**  *Provider and organizational willingness to deliver STI POCT* | | | | | |
| 1. To what extent were facilities and providers willing to adopt aetiological STI testing? 2. What are the main barriers/ facilitators to adopting the intervention? 3. What systems need to be in place for the health system to adopt intervention? | N/A | - Provider attitudes and motivations toward STI POCT - Compatibility with ANC workflows and resources - Organizational support - Views on training needs | Characteristics of Individuals | - Strong provider willingness to adopt STI POCT - Enhanced professional identity and credibility - Motivated by skill development and technology use | - *“Obviously it’s something interesting and a new technology and it will be beneficial for the patients… we are always eager to learn something new."* – (Professional Nurse, Facility A) |
|  |  |  | Intervention characteristics | - Perceived relative advantage of aetiological STI testing over syndromic management - Confidence in diagnostic accuracy - Usability of the GeneXpert platform - Acceptance of provider-collected vaginal swaps | - *“If a person tests positive for Gonorrhoea, we give direct treatment aimed at treating Gonorrhoea instead of the syndromic approach, which increases antimicrobial resistance.”* – (Research Nurse 2) - *“Simple is best. I don’t think it should cause any problem… negative is negative, positive is positive.”* – (Professional Nurse, Facility A) |
|  |  |  | Inner Setting | - Strong organisational acceptance of aetiological STI testing - Adoption dependent on resources, support, and cross-cadre training - Workload and patient volume as anticipated barriers to scale-up | - *“With the proper resources we’ll be able to carry out new technology and adapt to new ways to deal with STIs.”* – (Professional Nurse, Facility B) - *“There might be slight resistance, because of the added responsibility.”* – (Professional Nurse, Facility D) |
| **Implementation**  *Fidelity, feasibility, and delivery of STI POCT* | | | | | |
| 1. How did implementation vary by facility context and population characteristics? 2. What support and tools are needed for consistent delivery of the intervention? 3. What were the major changes/adaptations needed during implementation | - Overall treatment coverage and same-day treatment rates - Facility-level variation in treatment rates - Attendance Rates - Facility-level variation in follow-up rates | - Barriers and facilitators to waiting for results and attending follow-up visits (e.g., work, hunger, travel, childcare) - Experiences with workflow integration, staffing, infrastructure, and supply chains - Description of site-specific adaptations | Intervention Characteristics | - Compact size viewed as facilitator to space constraints - Unreliable electricity undermines operational feasibility - Cold-chain capacity insufficient for high-volume samples or strict refrigeration requirements - Prolonged assay turnaround time incompatibility in high-volume and rural clinics | - *“…in terms of making needed refrigeration, there could be a challenge that it will need something going into the facility.”* – (Professional Nurse, Facility C) - *“Government patients already have long waiting times… and then having to wait 90 minutes is too much.”* – (Professional Nurse, Facility D) - *“Yhoo! Wait for an hour? Never.”* – (Participant 5) |
|  |  |  | Outer Setting | - Socioeconomic and geographic constraints drive loss to follow-up - Catchment characteristics shape implementation feasibility | - *“Most of them are from rural areas and it’s a broad facility that caters for a large catchment area, so should we have positive results, the patient is not coming back. Even now, they jump from space to space.”* – (Professional Nurse, Facility D) - *“The clinic is in a bush veld… separate from residential areas. When participants leave and walk home, they get mugged.”* – Field Officer 2 |
|  |  |  | Inner Setting | - Substantial facility-level variation in implementation capacity - Unstable supply chains undermine implementation fidelity - Facility readiness determines consistent delivery | - *“Space is a challenge. Our facility is organized in a restricted manner.”* (Professional Nurse, Facility A) - “*If it’s out of stock then it’s out of stock… now we don’t have the rapid test for syphilis…we don’t know what’s happening”* (Professional Nurse, Facility C) |
|  |  |  | Process | - Enhanced tracing and follow-up strategies used during the trial are resource-intensive and unsustainable under routine care - Community Health Workers perceived as an effective strategy to support follow-up and tracing given adequate resources. | - *“At [Facility C] we are using the community health workers, if we can’t get the participant over the phone, we ask the CHWs.”* – (Research Nurse) - *“We have community health workers for tracing, but it is currently a problem for the patients that we are seeing.. not only pregnant women, but also the ones coming for chronic treatment and those that are defaulting and we need to trace them. We have a huge number of such patients and it's difficult to trace them, so it would be an additional challenge should we have another group that we would need to trace back for the community workers.”* – (Professional Nurse, Facility A) |
| **Maintenance**  *Perceived sustainability and integration into routine ANC* | | | | | |
| 1. What resources and system conditions are required for sustained delivery? 2. What adaptions are needed to integrate intervention into current practices? | N/A | - Provider and manager perspectives on long-term feasibility, cost, and accountability - Perceived sustainability within existing staffing, infrastructure, and procurement systems - Alignment with routine ANC services and potential for integration into other service touchpoints (e.g., family planning, ART, well-baby care) | Intervention Characteristics | - Uncertainty about financial sustainability - Concerns about accountability for procurement and maintenance | - *“It’s financial... Who pays for it? Who maintains it? The Department won’t be able to. We are struggling with small operational issues”* – (Nurse Manager, Facility B) - *“Maintenance is another challenge… The Department has a big challenge when it comes to the budget.”* – (Nurse Manager, Facility D) |
|  |  |  | Outer Setting | - Sociocultural and relationship factors risk reinfection and undermine sustained impact | - *“You don’t know what the man is up to, even as we speak I don’t know where he is or what he is doing you see..”* – (Participant 17) - *“I saw that my husband is having a foot that is walking sideline”* – (Participant 1) |
|  |  |  | Inner Setting | - Staff shortages and high turnover risks lack of trained human resources for sustained and consistent delivery. | - *“Currently we would not be able to provide any more services.. If there were to be anything else added, it would suffer should the proper human resources not be provided. Currently, other programs are suffering already so it could not happen here.”* – (Professional Nurse, Facility D) |
|  |  |  | Process | - Universal screening supports prevention but challenges capacity at scale at high-volume sites - Integration into routine service touchpoints is essential for sustainability - Bundled service delivery improves retention - Extending STI POCT beyond pregnancy may support long-term maintenance - Early and repeat screening are valued but limited by follow-up constraints | - *“With pregnant women… I think it’s better to assess and test everyone”* – (Professional Nurse, Facility B) - *“If everyone was to be tested, I don’t think that would be feasible at all… if we would have a screening tool to screen those ANC clients and then you can see who actually need to be tested.”* – (Professional Nurse, Facility A) - *“On the first visit you might test negative, but then the next visit the boyfriend comes back and no condom is used. So—every visit.”* – (Professional Nurse, Facility D) - *“What encouraged me [to return] is because I care about my health; I care about my baby’s health… that’s it. When you are pregnant, you are more vulnerable to STIs… that’s what pushed me to come, and I wanted to have a healthy pregnancy without any complications.”* – (Participant 8) |
