## Supplemental File 2 for "Implementation of point-of-care screening for *Chlamydia trachomatis, Neisseria gonorrhoeae*, and *Trichomonas vaginalis* among pregnant women in South Africa: a mixed-methods process evaluation of the Philani Ndiphile trial"

**Introductions:** Hello, my name is ______________________. I am a research assistant and on behalf of the Foundation for Professional Development, I would like to interview you as part of a research project/study to understand a bit more about your experiences in the Philani Study and your views on testing for sexually transmitted infections (STIs) during pregnancy. I hope we can speak casually. Please know that there are no right or wrong answers. Please take your time to respond as clearly and with as much detail as possible. What you say will not be shared or your personal information will not be shared with anyone outside of the research/study team, including your health providers (doctors, nurses). We really appreciate your time and openness in answering our questions. Your knowledge, experiences and insights will be extremely helpful.

This should take approximately 30-60 minutes of your time today.

In reference to the consent form which you have read/has been read, and already signed, do you have any questions before we begin?

___ ___ ___ ___	                  __________________     	___ ___ ___ ___

Participant ID (PTID)	                  Staff Initials	 	   	Date of Interview

| **INTERVIEWERS:** *Before the interview, please ask the participant the following questions. Similar to the rest of the interview, this information will remain confidential and will not be linked to the participant’s name in any way or shared with anyone outside the research team.* | | |
| --- | --- | --- |
| **Demographics for Philani participants** | | |
| 1. | What is your age? | oo Years |
| 2. | Where did you attend antenatal care? *(if participant is unclear, can name the clinics)* | o Ndevana clinic  o Grey gateway clinic  o Nontyatyambo clinic  o Empilweni Gompo clinic  o Duncan Village day hospital [DVDH] |
| 3. | Gestational age when enrolled in Philani | oo weeks [could confirm on REDCap] |
| 8. | How many times were you tested for STIs? | __ __ number |
| 9. | Were you treated for an STI? | o Yes  o No  o Prefer not to answer |
| 10. | **[If they were positive]** Which infection did the nurse give you treatment for? *(If they don’t remember/recall, tick don’t know)* | o Chlamydia trachomatis  o Neisseria gonorrhoeae  o Trichomonas vaginalis  o Don’t know  o Prefer not to answer |

**Icebreaker**

**Interviewer:** I would like to first get to know you a little better and learn more about your experience during your pregnancy.

1. To start off, how are you doing today?
2. Tell me a little bit about yourself.
3. Please tell me about the type of care you received from this facility during your pregnancy (*probe why the participant describes it as good or not).*

How often did you come to this facility for your pregnancy?

**I would now like to talk about testing for STIs during antenatal care (ANC).**

4. Please tell me about the information you were given about sexually transmitted infections also known as STIs.
   a. How does this compare to what you knew about STIs before (coming here)?
5. What made you seek care for these types of infections? *(probe for reasons)*
   a. Did you seek care for any STI symptoms before?
      i. Please describe the services that you received from the facility for these symptoms.
   b. How did you hear about our STI services?
      i. How did you feel about the information you were given?
   c. In your own words, can you describe what happened on the day you were first tested for STIs?
      i. What did you like about this visit? *(probe: why they felt comfortable or not)*
      ii. What could have been different during this visit?
      iii. Did you feel that anything was missing?
      iv. What happened after?
6. How would you feel about the following. Imagine you were asked to come back to the clinic for a separate test 21 days after you tested STI-positive, to check whether your infection has been cured?

*a. (Probe for reasons if willing to test or not at 21 days)*

7. Please describe how you would feel about testing for STIs again at the routine 32 week (8 months) ANC visit?
   a. *(Probe for reasons if willing to test or not at 32 weeks)*
8. When would you prefer to be tested for STIs? (*Note for interviewer: can be any time*)
   a. How often would you prefer to be tested for STIs?
   b. Would this be different if you were pregnant or not? Why?
9. You mentioned earlier you were tested X [number] of times at this facility. How did you feel about the number of times you were tested for STIs?
   a. How many times were you asked to come to the site for testing?
      i. Please describe how you received information from staff about your follow-up visits?
         1. How did you feel about this?
   b. What encouraged you to return for these tests?
      i. How does being pregnant or not influence returning for STI testing?
   c. What challenges did you experience? *(e.g., returning, testing)*
   d. What would you have preferred?

**Now we will talk a bit more about your experiences during the Philani study and what it was like waiting for your results.**

10. In your own words, what was it like to be tested for STIs?
    a. How did you feel about the way that specimens were collected? (i.e., vaginal swabs)
       i. How did the staff explain this to you?
       ii. How did you feel about the number of specimens that were collected?
       iii. How do you prefer specimens to be collected?
            1. Why do you prefer this method?
    b. What did you think when you saw the GeneXpert machine (the device used to test you for STIs)?
       i. What did the staff tell you about the machine?
11. On the day you were tested, you would’ve noticed you had to wait for 90 minutes for your test results. Could you describe what happened while you were waiting for your results?
    a. Were you able to wait for your test results on the same day?
       i. **(If yes..)** Please describe where you were waiting for the results.
          1. Did you experience any challenges during this time?
          2. Please describe what happened at the clinic after your results were known.
       ii. **(If no…)** What were some of the challenges you faced having to wait for your results?
           1. Would you wait this amount of time for any other types of tests?
           2. What challenges do you face in returning to the clinic later to obtain your test result or treatment (if needed)?
           3. What encouraged you to return for your results (if applicable)?
    b. Would you normally be willing to wait ~90 minutes at the clinic to obtain your STI test results?
       i. **[If no]** Could you please describe what the challenge is with waiting this long for your test result at the clinic?
          1. Would you be willing to come back to the clinic another time/day to receive your results? *(Probe why or why not)*
          2. When would you be willing to come back to the clinic for your next visit? *(such as the next day, next week, or the next scheduled visit)?*
       ii. **[if yes]** Why would you normally be willing to wait ~90 minutes?
12. If you could get STI treatment right away after your test results, how would that affect your willingness to wait about 90 minutes for those results?
    a. [**If still unwilling to wait…]** How could STI testing be done differently?
13. Please describe any challenges you experienced relating to the STI care you received?
    a. What barriers/challenges did you face taking or completing your treatment?
14. What encouraged you to take your treatment?
15. What do you think can be improved when it comes to screening for STIs?
    a. What could we change at the healthcare facilities when it comes to STIs?

**I want to now understand what impact testing for STIs had for you and if you have any recommendations.**

16. Please describe how testing and receiving treatment for STIs benefited you?
17. Did you seek additional/other care for STIs outside of this study? Could you please give an example?
18. In the future, where would you seek care for STIs or where would you go if you had concerns about STIs?
    a. Why would you go there?
    b. What type of services would you ask for in the future?
19. How would you prefer to be tested for STIs? *Probe: self-versus nurse collected swabs, type of test, location, timing of testing (e.g., during antenatal care, any visit, medication pick-up, family planning)*
    a. When would be the best time to test for STIs? (*during pregnancy, after delivery, during family planning etc*)
    b. Who would you prefer administers these types of tests?
    c. If new tests were being developed for STIs, what things are important to consider as part of this test? (*e.g., timing, technical features, accuracy*)
20. Do you have any recommendations or suggestions based on your experiences with testing?

**Introductions:** Hello, my name is ______________________. I am a research assistant and on behalf of the Foundation for Professional Development, I would like to ask you some questions to better understand your views on testing for sexually transmitted infections (STIs) and the need for STI screening within the Eastern Cape.

In reference to the consent form which you have read/has been read to you, and already signed, do you have any questions before we begin?

**Demographic form**

___ ___ ___ ___	                          __________________	     	___ ___ ___ ___

Participant ID (PTID)	                  Staff Initials	 	   	              Date of Interview

| **INTERVIEWERS:** Before the interview, please ask the participant the following questions. Similar to the rest of the interview, this information will remain confidential and will not be linked to the participant’s name in any way or shared with anyone outside the research team. | | |
| --- | --- | --- |
| **For key stakeholders including providers, NDOH staff and research staff** | | |
| 1. | What is your occupation? | ☐_1_ |
| 2. | Years in current occupation | oo Years |
| 3. | Affiliation | o NDOH  o NHLS  o FPD  o Other: ____________ |
| 4. | How long have you worked for your organization? | oo Years |
| 5. | What is your gender? |  |
| 6. | How old are you? | oo Years |
| 7. | What role do you play in STI screening? |  |
| 8. | What other aspects of clinical care are you involved in? |  |

**Role and perception of STIs and POCT**

1. Can you describe your specific role in STI screening, diagnosis, and care for pregnant women?
   a. How does this role align with your overall responsibilities in antenatal care (ANC)?
2. What challenges do you currently face in diagnosing and treating STIs in pregnant women?
   a. How do you think point-of-care testing (POCT) could address these gaps?
3. How do you think the implementation of STI POCT aligns with broader maternal and newborn health goals at your facility?

**Barriers and Facilitators to POCT Implementation**

4. What are the key challenges or barriers to adopting STI POCT at your facility? *[Probe: resource-related, structural, staff attitudes/perceptions]*
5. What factors or conditions at your facility would enable the successful implementation of STI POCT?
   a. Are there existing practices, resources, or support systems that could help facilitate this process?
6. Do you feel there is adequate organizational support to implement STI POCT in ANC? *[Probe: why or why not]*
   a. What support from facility management or the health system would be critical for the successful implementation of POCT for STIs?

**Implementation considerations for STI POCT**

7. What infrastructure improvements would be necessary to support the implementation of POCT for STIs at your facility?
   a. What infrastructure improvements would be necessary to support the implementation of POCT for STIs at your facility?
8. Which staff members at your facility would likely be responsible for conducting STI POCT for pregnant women?
   a. Do you think the current workforce is adequately staffed to take on this responsibility?
   b. What type of training or capacity-building would staff need to effectively implement STI POCT?
9. What implications would STI POCT have on laboratory services at your facility? *[Probe: limitations in specimen storage space, testing capacity, staff resources]*
10. Do you think STI POCT complements the existing ANC service delivery workflow?
    a. How could STI POCT be effectively integrated with other ANC services (e.g., HIV and syphilis testing, ultrasound).
11. Given resource constraints, do you believe targeting STI POCT to high-risk women, such as those living with HIV, is a feasible approach at your facility?
    a. What existing strategies or modalities support targeted testing or interventions for high-risk pregnant women at your facility?
    b. **If no** **strategies exist,** what resources or approaches would be necessary to implement targeted testing?

**Timing and frequency of STI screening:**

12. At what point(s) during pregnancy do you think STI screening should be conducted?
13. How frequently do you believe STI screening should occur during pregnancy?
    a. Do you believe this frequency applies to all pregnant women, or are there specific groups who should be screened more often?
14. Is there value in conducting repeat STI POCT at the 32-week ANC visit for all pregnant women? [Probe: why or why not]
    a. How could repeat third trimester STI POCT be effectively integrated with existing testing schedules or services at this timepoint? (e.g., HIV and syphilis testing)
15. Should pregnant women who test positive for STIs be required to return to the clinic to receive a test of cure 3 weeks after treatment? *[Probe: why or why not]*
    a. Do you anticipate challenges or barriers for women to return to the clinic for a non-routine visit?
    b. **If yes,** how might these challenges be addressed?

**STI Test Results**

16. How are point-of-care and laboratory test results currently reported back to pregnant women at your facility?
    a. Do you think STI test results should be reported back in the same way? *[Probe: why or why not].*
    b. **If not**, what alternative methods do you think would work better for reporting STI test results?
17. Do you think POCT for STIs could influence the process of notifying and encouraging partners of pregnant women to get tested and treated for STIs? *[Probe: why or why not].*
    a. Are there specific strategies you believe could be implemented alongside POCT to improve partner testing rates?
    b. What challenges might still exist in ensuring partners are treated promptly, even with POCT?

**POCT turnaround time**

18. Considering the current turnaround time for STI POCT is approximately 90 minutes, are there areas at your facility where women can wait for their test results?
    a. **If such areas exist**, can you describe the infrastructure?
    b. **If no such areas exist**, what infrastructure changes would be necessary to enable women to wait for their results?
    c. What strategies could health care workers use to encourage women to wait for their results?
19. What are the biggest challenges or barriers to following up with women who leave without receiving their test results?
    a. How might these barriers be addressed with additional resources or process changes?
20. If a woman does not wait for her test results, what strategies or opportunities exist to ensure she receives her test results and appropriate treatment?
    a. Do you think your facility or community has the resources (e.g., staff, time, technology) to implement these follow-up strategies effectively?
21. What communication methods (e.g., phone calls, SMS, home visits) are currently used at your facility to follow up with women who miss appointments or test results?
    a. Could these methods be adapted or expanded for STI POCT results?
22. Are there existing community health worker programs or community-based initiatives that could assist in following up with women who leave without their test results?
    a. How could these programs be leveraged for STI POCT follow-up specifically?
23. Does your facility use any patient tracking or record-keeping systems to monitor women who do not return for test results?
    a. **If yes**, how effective are these systems, and how might they be improved to support follow-up for STI POCT results?
    b. **If no**, what type of system or tool would be most helpful for tracking and follow-up?

**Health care worker engagement:**

24. What approaches would ensure healthcare workers are fully engaged and motivated to adopt POCT for STIs into their practice?
25. How can concerns or resistance from providers about implementing POCT be addressed?
26. If new tests were being developed for diagnosing STIs, what features are important to consider as part of this test? (e.g., timing, technical features, accuracy)
27. Do you have any recommendations or suggestions based on your experiences with screening pregnant women for STIs?

**Introductions:** Hello, my name is ______________________. I am a research assistant and on behalf of the Foundation for Professional Development I would like to ask you some questions to better understand your views on testing for sexually transmitted infections and the need for STI screening within the Eastern Cape. Please know that there are no right or wrong answers. Please take your time to respond as clearly and with as much detail as possible. What you say will not be shared or your personal information will not be shared with anyone outside of the qualitative research/study team. We really appreciate your time and openness in answering our questions. Your knowledge, experiences and insights will be extremely helpful. This should take approximately 30-60 minutes of your time today.

In reference to the consent form which you have read/has been read to you, and already signed, do you have any questions before we begin?

**Demographic form**

___ ___ ___ ___	                          __________________	     	___ ___ ___ ___

Participant ID (PTID)	                  Staff Initials	 	   	              Date of Interview

| **INTERVIEWERS:** *Before the interview, please ask the participant the following questions. Similar to the rest of the interview, this information will remain confidential and will not be linked to the participant’s name in any way or shared with anyone outside the research team.* | | |
| --- | --- | --- |
| **For key stakeholders including providers, NDOH staff and research staff** | | |
| 1. | What is your occupation? | ☐_1_ |
| 2. | Years in current occupation | 🞏🞏 Years |
| 3. | Affiliation | 🞏 NDOH  🞏 NHLS  🞏 FPD  🞏 Other: ____________ |
| 4. | How long have you worked for your organization? | 🞏🞏 Years |
| 5. | What is your gender? |  |
| 6. | How old are you? | 🞏🞏 Years |
| 7. | What role do you play in STI screening? |  |
| 8. | What other aspects of clinical care are you involved in? |  |

**First of all, I’d like to talk about your role, and how you integrate STI testing in other services.**

1. What is your role in STI screening and care?
   a. Tell me a bit more about your activities and responsibilities related to STIs
   b. What do you find interesting about STIs?
      i. What do you think are current challenges?
2. What was interesting about your work providing STI screening services using Xpert?
   a. Do other staff share this interest? *(Probe why/why not)*
3. How did you create organizational support at your facility for the delivery of STI testing to pregnant women?

**We would like to find out more about the experience implementing the Philani study and screening pregnant women for STIs.**

4. How much time is needed to screen a patient for STIs using point-of-care tests (POCT)?
   a. Could you briefly describe the flow of STI screening at your facility
   b. What impacts the duration or the flow of screening for STIs?
      i. Please provide examples.
5. What are the barriers and enablers that impacted the implementation of Philani?
   c. What challenges did you experience using POCT to screen pregnant women for STIs?
      i. How were these resolved?
   d. What challenges did you experience implementing the TOC for women 3 weeks following STI treatment?
      i. How were these resolved?
   e. What challenges did you experience implementing the repeat POCT at the 32 week visit?
      i. How were these resolved?
6. How was Philani adapted over time at your facility from the original protocol?
   f. Please describe the strategies that were used
   g. What were the additional resources that were required
7. Does STI POC complement existing services at the facility? *(Why, why not)*
8. What challenges did you face in reaching women who did not wait to provide their test result?
9. What challenges did you face in delivering STI treatment to women who did not wait for their test results?
10. For women who did not wait for the POCT results, what strategies did you employ to reach them and provide them with treatments?
    a. How long did this typically take?

**I would like to ask you a few questions about the adoption and any implementation recommendations for STI screening at primary healthcare level.**

11. What organizational change would be needed for STI POCT for your work setting?
12. How do you think STI screening may be adapted going forward based on your experiences?
    a. When was the optimal time to screen women for STIs during pregnancy?
    b. Was there an optimal time to retest pregnant women for STIs during pregnancy? [Why do you say so?]
    c. Was there an optimal time to conduct a test of cure for women following treatment? [Why do you say so?]
    d. What adaptions might be needed to prioritize women at increased risk of baseline or incident STIs, given limited resources?
13. How has this study changed STI care/services for pregnant women in your study setting?
14. Did you have the resources to adequately perform STI POC and what were these?
15. What are crucial factors for the sustainability of POC testing and treatment going forward?
16. Do you have any other recommendations for how we could think differently about screening for STIs?
17. We have come to the end of our interview, is there anything else you’d like to discuss/add?

Thank you very much for your time, your feedback will be very helpful.
