## Supplemental File 4 for "Implementation of point-of-care screening for *Chlamydia trachomatis, Neisseria gonorrhoeae*, and *Trichomonas vaginalis* among pregnant women in South Africa: a mixed-methods process evaluation of the Philani Ndiphile trial"

### Good Reporting of A Mixed Methods Study (GRAMMS)

| Guideline | Section: page |
| --- | --- |
| Describe the justification for using a mixed methods approach to the research question | Process Evaluation and Theoretical Framework p8 |
| Describe the design in terms of the purpose, priority and sequence of methods | Process Evaluation and Theoretical Framework p8 |
| Describe each method in terms of sampling, data collection and analysis | Data collection p8-9<br>Data analysis p10 |
| Describe where integration has occurred, how it has occurred and who has participated in it | Data analysis p10 |
| Describe any limitation of one method associated with the present of the other method | Discussion p29 |
| Describe any insights gained from mixing or integrating methods | Results 14-24<br>Discussion 25-29 |

*O'Cathain A, Murphy E, Nicholl J. The quality of mixed methods studies in health services research. J Health Serv Res Policy. 2008;13(2):92-98.*
