## Supplemental File 5 for "Implementation of point-of-care screening for *Chlamydia trachomatis, Neisseria gonorrhoeae*, and *Trichomonas vaginalis* among pregnant women in South Africa: a mixed-methods process evaluation of the Philani Ndiphile trial"

### BMJ Global Health Author Reflexivity Statement

Adapted from Morton, B., Vercueil, A., Masekela, R., Heinz, E., Reimer, L., Saleh, S., Kalinga, C., Seekles, M., Biccard, B., Chakaya, J., Abimbola, S., Obasi, A. and Oriyo, N. (2022), Consensus statement on measures to promote equitable authorship in the publication of research from international partnerships. Anaesthesia, 77: 264-276. <https://doi.org/10.1111/anae.15597>

| **Study conceptualisation** | |
| --- | --- |
| 1. How does this study address local research and policy priorities? | This study addresses South African and provincial priorities to reduce adverse maternal and neonatal outcomes and strengthen STI and HIV prevention by generating implementation evidence to inform integration of POC STI screening into routine antenatal care. |
| 2. How were local researchers involved in study design? | Researchers based at the University of Cape Town and local implementing partner (Foundation for Professional Development) were centrally involved in study design, ensuring alignment with national guidelines, clinic workflows, and health system priorities in the Eastern Cape. |
| **Research management** | |
| 3. How has funding been used to support the local research team(s)? | Funding supported local research nurses, fieldworkers, coordinators, and data teams, as well as training, site operations, and implementation of clinic-based research activities. |
| **Data acquisition and analysis** | |
| 4. How are research staff who conducted data collection acknowledged? | Local research staff are acknowledged in the manuscript and supplementary materials, with key contributors included as co-authors where appropriate. |
| 5. How have members of the research partnership been provided with access to study data? | Data were managed through secure platforms and shared with authorised partners at UCT and collaborating institutions in accordance with ethical approvals and data governance procedures. |
| 6. How were data used to develop analytical skills within the partnership? | Local researchers were actively involved in quantitative and qualitative analysis, including use of R and NVivo, with mentorship and collaborative analysis processes. |
| **Data interpretation** | |
| 7. How have research partners collaborated in interpreting study data? | Data interpretation was conducted collaboratively through regular meetings involving UCT investigators, local research teams, and implementation partners. |
| **Drafting and revising for intellectual content** | |
| 8. How were research partners supported to develop writing skills? | Researchers were supported through mentorship, iterative feedback, and active involvement in drafting and revising the manuscript. |
| 9. How will research products be shared to address local needs? | Findings will be disseminated to participating clinics, the Eastern Cape Department of Health, and national stakeholders through reports, presentations, and policy engagement. |
| **Authorship** | |
| 10. How is the leadership, contribution and ownership of this work by LMIC researchers recognised within the authorship? | LMIC researchers, including those based at UCT and South African institutions, hold lead and senior authorship positions, reflecting their central role in study design, implementation, analysis, and interpretation. |
| 11. How have early career researchers across the partnership been included within the authorship team? | Early career researchers were actively involved in primary and supportive data collection, analysis, and manuscript development, with inclusion in authorship based on contribution. |
| 12. How has gender balance been addressed within the authorship? | The authorship team reflects gender diversity, with strong representation of women across early researcher and senior roles. |
| **Training** | |
| 13. How has the project contributed to training of LMIC researchers? | The project provided hands-on training in implementation research, process evaluation analysis, and qualitative methods for local researchers and research staff. |
| **Infrastructure** | |
| 14. How has the project contributed to improvements in local infrastructure? | The study supported clinic-based research infrastructure, including deployment of diagnostic platforms, strengthening of data systems, and integration of research activities within routine ANC workflows |
| **Governance** | |
| 15. What safeguarding procedures were used to protect local study participants and researchers? | Ethical approval was obtained from the University of Cape Town Human Research Ethics Committee. Written informed consent was obtained from all participants, and procedures ensured confidentiality, voluntary participation, and protection of both participants and research staff. |
