## Supplemental File 6 for "Implementation of point-of-care screening for *Chlamydia trachomatis, Neisseria gonorrhoeae*, and *Trichomonas vaginalis* among pregnant women in South Africa: a mixed-methods process evaluation of the Philani Ndiphile trial"

**Supplemental File 6: Characteristics of Interview and Focus Group Discussion Participants**

| Individual Interview Participants | **N = 20**^1^ |
| --- | --- |
| Age at enrolment (years) | 30 (23.5, 31.25) |
| Gestational age at enrolment (weeks) | 11 (8, 16) |
| Facility |  |
| A (Urban) | 10 (50%) |
| B (Peri-urban) | 4 (20%) |
| C (Rural) | 4 (20%) |
| D (Peri-urban) | 2 (10%) |
| Philani Ndiphile Research Arm |  |
| Intervention Arm 1 | 9 (45%) |
| Intervention Arm 2 | 11 (55%) |
| Waited for POC STI test results at baseline visit |  |
| Yes | 11 (55%) |
| No | 9 (45%) |
| Positive for any STI* at baseline |  |
| Yes | 10 (50%) |
| No | 10 (50%) |
| Focus Group Discussion Participants | **N = 21**^1^ |
| Stakeholder group |  |
| Philani Ndiphile Study Staff | 7 (33.3%) |
| Facility-Based Healthcare Workers | 14 (66.6%) |
| Gender |  |
| Male | 4 (19%) |
| Female | 17 (81%) |
| Profession |  |
| Research Nurse | 3 (14.3%) |
| Research Field Worker | 4 (19%) |
| Professional Nurse | 11 (52.3) |
| Nurse Practitioner | 1 (4.8%) |
| Nurse Manager | 2 (9.5%) |
| Facility± |  |
| A (Urban) | 5 (35.7%) |
| B (Peri-urban) | 5 (35.7%) |
| C (Rural) | 2 (14.3%) |
| D (Peri-urban) | 2 (14.3%) |
| ^1^n (%); Median (Q1, Q3)  * Includes *Chlamydia trachomatis*, *Neisseria gonorrhoea*, or *Trichomonas virginals*  ± Only reported among facility-based healthcare workers (n = 14) | |
